# Adverse events associated with nifurtimox treatment for Chagas disease in children and adults

**DOI:** 10.1101/2020.06.02.20118000

**Authors:** A.J. Berenstein, N. Falk, G. Moscatelli, S. Moroni, N. González, F. Garcia-Bournissen, G. Ballering, H. Freilij, J. Altcheh

**Affiliations:** Instituto Multidisciplinario de Investigaciones en Patologías Pediátricas (IMIPP), CONICET-GCBA, Laboratorio de Biología Molecular, División Patología, Hospital de Niños Ricardo Gutiérrez, Buenos Aires, Argentina; Parasitología, Hospital de Niños Ricardo Gutierrez and Instituto multidisciplinario de Investigación en Patologías Pediátricas -(IMIPP) CONICET-GCBA, Buenos Aires, Argentina; Division of Paediatric Clinical Pharmacology, Department of Paediatrics, Schulich School of Medicine and Dentistry, Western University. London, Ontario, Canada

**Author notes:** Both authors contributed equally (A.J. Berenstein, N. Falk).

**Keywords:** Trypanosoma cruzi, nifurtimox, children, adults, adverse drug reactions

## Abstract

**BACKGROUND:** Nifurtimox (NF) is one of the only two drugs currently available for Chagas disease (ChD) treatment. However, there is scarce data on NF safety, and many physicians defer or refuse NF treatment because of concerns about drug tolerance.

**METHODS:** Prospective cohort study with retrospective data collection of adverse drug reactions (ADRs) associated with NF treatment of ChD. Children received NF doses of 10-15 mg/kg/day for 60-90 days, and adults 8-10 mg/kg/day for 30 days.

**RESULTS:** 215 children (median age: 2.6yrs, range 0-17) and 105 adults (median age: 34yrs, range 18-57) were enrolled. Overall, 127/320 (39.7%) patients developed ADRs, with an incidence of 64/105 in adults, and 63/215 in children (OR = 3.7, 95%CI [2.2;6.3]). We observed 215 ADRs, 131 in adults (median: 2 events/patient (IQR_25-75_= 1-3) and 84 in children (median: 1 event/patient (IQR_25-75_= 1-1.5) (Wilcoxon-Test, P_Adjusted_ < 8.10^-5^). ADRs were mainly mild and moderate. Severe ADRs were infrequent (1.2% in children and 0.9% in adults). Nutritional, central nervous and digestive systems were the most frequently affected, without differences between both groups.

Treatment was discontinued in 31/320 (9.7%) patients without differences between groups. However, ADR-related discontinuations occurred more frequently in adults than in children (OR = 5.5, 95%CI = [1.5;24]).

**CONCLUSIONS:** Our study supports the safety of NF for ChD treatment. Delaying NF treatment due to safety concerns does not seem to be supported by the evidence.

## Background

Chagas disease (ChD) is a silent but devastating disease caused by infection with the parasite *Trypanosoma cruzi*. The disease is endemic to the Americas, from the USA to Argentina, with over 7 million people currently infected in Latin America. ChD has expanded to many countries of the world via immigration, most cases reported in Europe, North America, Australia and Japan[1].

Most patients are asymptomatic during acute ChD. The acute phase is followed by a chronic asymptomatic stage that will eventually lead to irreversible heart disease in up to 30% of the infected patients many years later[2]. Over 7,000 deaths occur yearly due to complications of Chagas.

The current treatment for ChD is limited to two nitro-heterocyclic drugs, nifurtimox (NF) and benznidazole (BZ), both with similar effectiveness. Despite both drugs having been available since the early 70s, treatment recommendations vary significantly from country to country and the evidence-base for the current treatment regimens is limited. Regardless of treatment guidelines for ChD [3] that indicate treatment, many physicians and patients defer or refuse treatment because of anxiety over drug safety. According to previous studies, the most common side effects for both drugs were anorexia and weight loss, irritability, and less commonly, skin rashes [4,5].

As in BZ, the ADRs of NF are most commonly observed in adults, and children seem to tolerate NF treatment better[6]. However, current tolerability data comes mainly from small cohort studies. The pharmacological basis for the differences in the incidence of adverse events remain to be studied [7].

Here, we present results from a large cohort of ChD patients including infants, children and adults treated with NF, describing and comparing safety among adults and pediatric patients.

## Methods

### Study design and population

This is a prospective age-stratified cohort study with retrospective data collection, to assess safety and tolerability of oral NF in subjects with ChD. All patients were treated and followed-up at the Parasitology and Chagas service, Hospital de Niños “Ricardo Gutiérrez”, Buenos Aires, Argentina from January 1980 to July 2019.

Patients were stratified according to age. Sub-analysis among children was done considering the following age groups: (0 - 7mos), (8mos - 1yr), (2 - 6yrs), (7 - 17yrs).

**Chagas Disease diagnostic criteria:** For infants younger than 8 months: direct observation of *T.cruzi* using parasitological concentration method (microhematocrit test, MH) or xenodiagnosis (XD); for older patients: 2 reactive serological tests: Enzyme Linked Immunosorbent Assay (ELISA), Indirect Hemagglutination (IHA) or Direct agglutination (DA).

### Treatment

NF treatment (120-mg tablets, Bayer) was prescribed in doses of 10-15 mg per kg per day divided in two or three daily doses for 60 to 90 days for infants and children, and 8-10 mg per kg for 30 days for adults, according to national guidelines. Enrollment of children started in January 1980, and enrollment of adults started in July 2008. Infant NF doses were provided as fractionated tablets prepared by a pharmacist and administered with water or mother’s milk. Medication was provided to patients or their guardians in monthly batches, and compliance was assessed by counting remaining tablets at each visit. Treatment was considered complete when patients took the medication for at least 60 days for children and 30 days for adults.

### Data Collection

Data were collected from medical records of treated patients and entered into an Access clinical database (ACD) designed for this study. All individual datasets were anonymized. Demographic data, clinical and biochemical assessments and complementary studies were collected during follow-up. Baseline data values were obtained at the beginning of the treatment. Visits were carried out at 7, 30 days and at the end of treatment, every 3 months during the first-year post-treatment and every 6-12 months thereafter.

ADRs were evaluated through laboratory tests, clinical interviews and physical examinations, and classified according to World Health Organization (WHO) definitions [8,9]. Causality assessment was performed using the WHO criteria for causality assessment

Information on treatment duration and dosage, temporary interruptions and concomitant medications was systematically collected from medical records and documented in the clinical Database.

### Statistical Analysis

Continuous variables were expressed with mean and median, as applicable, with the corresponding standard deviation or interquartile range. Categorical variables were expressed in percentages. To test for significance, as appropriate, T test or Wilcoxon unpaired rank test (W.T.) for continuous variables and Fisher exact tests (F.E.T.) for categorical ones. P-values were adjusted by false discovery rate (Benjamini-Hochberg procedure). Adjusted p-values with P_Adjusted_< 0.05 were considered statistically significant. The statistical package R was used [10].

### Ethics statement

Study protocol was approved by the research & teaching committee and the bioethics committee of the Buenos Aires Children’s Hospital “Dr Ricardo Gutierrez”. The protocol was registered at ClinicalTrials.gov (NCT#04274101).

## Results

### Population characteristics

Medical records of ChD patients treated at our Institution were reviewed, and 372 patients who were prescribed NF were identified. However, 52 patients were excluded because they did not start treatment (i.e. did not fill in NF prescription). The remaining 320 patients were included in the cohort. A total of 215 pediatric patients [0-18yrs) and 105 adults were included. Among children were: n=56 (0 - 7mos) n=43 (8mos - 1 yr), n=44 (2 - 6yrs) and n=72 (7 - 17yrs).

In general, male and female subjects were well balanced in children but not in adults where 87.6% of subjects were female (most of them were mothers of children assisted in our service). The route of infection was: congenital in 131, undetermined in 154, vectorial in 32 and by blood transfusion in 3 cases (Table 1).

**Table 1.** Demographic data.

|  | Children (%) | Adults (%) | Total Patients (%) |
| --- | --- | --- | --- |
| **Gender** |  |  |  |
| F | 109 (50.7) | 92 (87.6) | 201 (62.8) |
| M | 106 (49.3) | 13 (12.4) | 119 (37.2) |
| **Age (months)** |  |  |  |
| Median [Q1, Q3] | 31.0 [6.00, 108] | 408 [348, 456] | 108 [13.8, 348] |
| Mean (SD) | 59.5 (63.1) | 414 (88.5) | 176 (182) |
| Min-Max | 1.00-215 | 228-684 | 1.00-684 |
| **Route of Infection** |  |  |  |
| Vector | 22 (10.2) | 10 (9.5) | 32 (10.0) |
| Congenital | 120 (55.8) | 11 (10.5%) | 131 (40.9) |
| Blood transfusion | 1 (0.5) | 2 (1.9) | 3 (0.9) |
| Undetermined | 72 (33.5) | 82 (78.1) | 154 (48.1) |
| **Clinical Examination at diagnosis** |  |  |  |
| Asymptomatic | 185(86.0) | 104(99.0) | 289(90.3) |
| Symptomatic | 30(14.0) | 1(1.0) | 31(9.7) |
| **Total** | **215(67)** | **105(33)** | **320(100)** |

F: Female, M: Male

Overall, most patients were asymptomatic 289/320 (90.3%). However, patients infected by the vector route were predominantly symptomatic 12/21 (57.1%) and the most frequent symptom was the ocular chagoma in 11/12 cases. In patients infected by the congenital and undetermined route, symptomatic cases were observed mainly in infants under 2 years 16/19 (84.2%), and the most affected organ was the liver (12/19 cases).

A clinical improvement was observed in 30/31(96.8%) of symptomatic patients during treatment. An infant in the range of 0-8 months, coinfected with *T.cruzi* and HIV by the transplacental route, developed encephalitis and myocarditis, and died during NF treatment due to respiratory complications.

### ADRS incidence and relationship

Overall, 127/320 patients (39.7%) developed ADRs, with an incidence in adults of 64/105 (60.9%), and of 63/215 (29.3%) in children (F.E.T., OR = 3.7, CI95% = [2.2 - 6.3]; P_Adjusted_ = 3.5 10^-7^).

A total of 215 ADRs in 127 patients were observed. In 64 adults, 131 ADRs were observed with a median of 2 events per patient (IQR_25-75_ = 1-3) and in 63 children, 84 ADRs were observed with a median of 1 event per patient (IQR_25-75_ = 1-1.5); (W.T. P_Adjusted_ = 7.9 10^-5^).

NF-related ADRs were more frequent in adults (79.7%) than in children (7.9%) (F.E.T., OR=9.9, CI95% = [3.7 - 33]; P_Adjusted_ = 1.3.10^-7^; see Table 2 for further details).

**Table 2.**
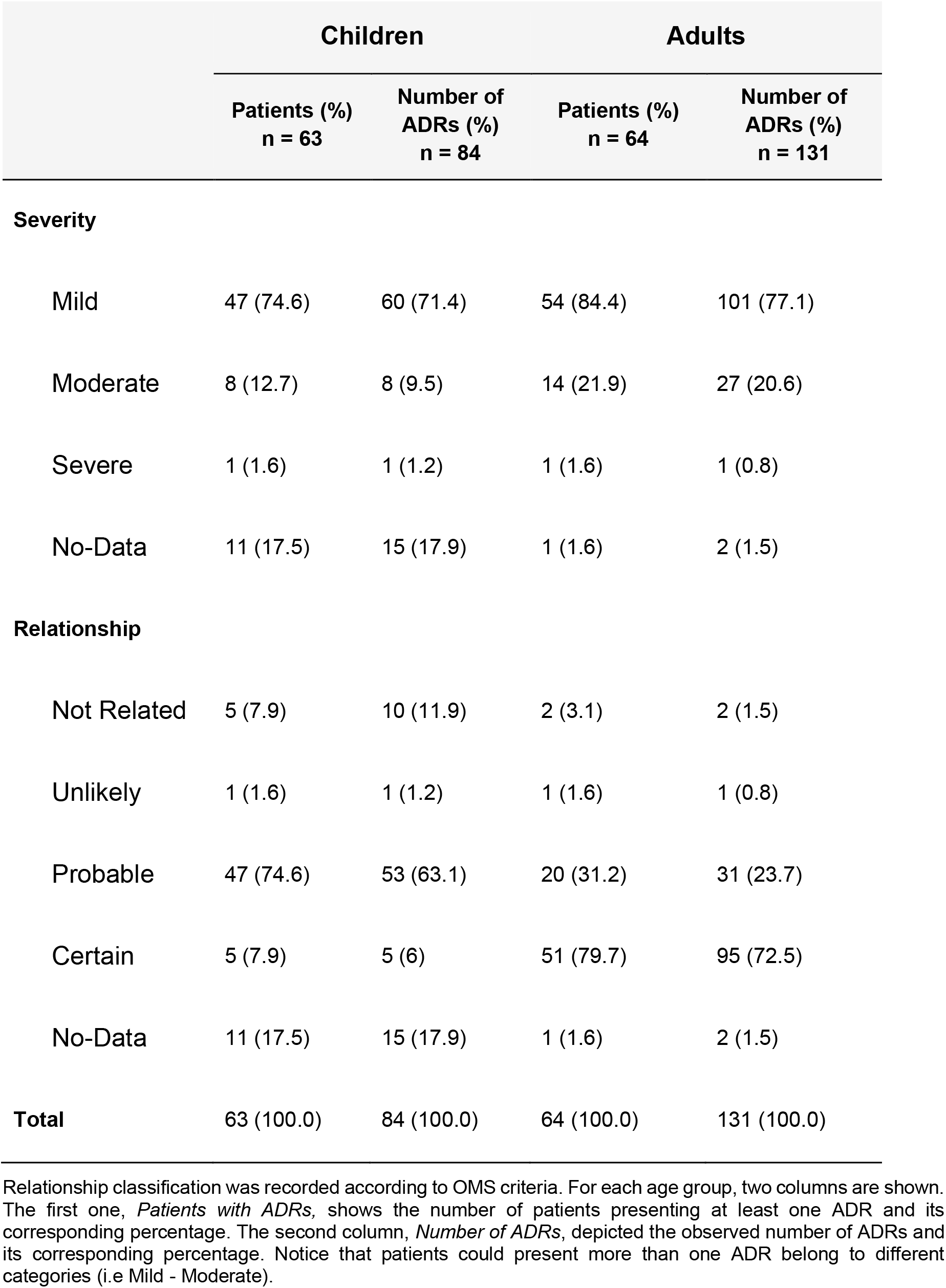
Adverse events classified by severity and their relationship to treatment

|  | Children |  | Adults |  |
| --- | --- | --- | --- | --- |
|  | Patients (%)<br>n = 63 | Number of<br>ADRs (%)<br>n = 84 | Patients (%)<br>n = 64 | Number of<br>ADRs (%)<br>n = 131 |
| **Severity** |  |  |  |  |
| Mild | 47 (74.6) | 60 (71.4) | 54 (84.4) | 101 (77.1) |
| Moderate | 8 (12.7) | 8 (9.5) | 14 (21.9) | 27 (20.6) |
| Severe | 1 (1.6) | 1 (1.2) | 1 (1.6) | 1 (0.8) |
| No-Data | 11 (17.5) | 15 (17.9) | 1 (1.6) | 2 (1.5) |
| **Relationship** |  |  |  |  |
| Not Related | 5 (7.9) | 10 (11.9) | 2 (3.1) | 2 (1.5) |
| Unlikely | 1 (1.6) | 1 (1.2) | 1 (1.6) | 1 (0.8) |
| Probable | 47 (74.6) | 53 (63.1) | 20 (31.2) | 31 (23.7) |
| Certain | 5 (7.9) | 5 (6) | 51 (79.7) | 95 (72.5) |
| No-Data | 11 (17.5) | 15 (17.9) | 1 (1.6) | 2 (1.5) |
| **Total** | 63 (100.0) | 84 (100.0) | 64 (100.0) | 131 (100.0) |

Relationship classification was recorded according to OMS criteria. For each age group, two columns are shown. The first one, *Patients with ADRs*, shows the number of patients presenting at least one ADR and its corresponding percentage. The second column, *Number of ADRs*, depicted the observed number of ADRs and its corresponding percentage. Notice that patients could present more than one ADR belong to different categories (i.e Mild - Moderate).

The number of ADRs was associated with incomplete treatment: 2.58 events/patient in subjects that discontinued treatment vs 1.55 events/patient in those with completed treatment (W.T., P_Adjusted_ = 1.6 10^-4^). Moreover, differences remained when considering both cohorts separately (adults: W.T. P_Adjusted_ = 0.028; children: W.T. P_Adjusted_ = 0.016.). The profile of ADRs is shown in Table 3. The systems most commonly affected were nutritional 75/215 (34.9%), Central Nervous System (CNS) 61/215 (28,4%) and digestive 38/215 (17.7%) without differences between adults and children. Few adverse skin effects (20/215, 9.3%) were observed in both groups and hematological ADRs (7/215 events) were observed only in children (F.E.T., OR = Inf, CI95% = [2.3 - Inf]; *P*_Adjusted_=0.0047). Time of onset of ADRs was recorded for 130/215 ADRs in 64/127 patients (50.4%). Overall, 93.8% of ADRs appeared within 30 days of treatment. ADRs median onset time (IQR_25-75_) was: 5 (2.5 - 10.2) days for digestive, 5.5 (1.5 - 11) days for CNS, 8 (0.75 - 26) days for nutritional, and 13 (10 - 20) days for skin.

**Table 3.** ADR occurrence and patient incidence by organ system.

|  | Children |  | Adults |  |
| --- | --- | --- | --- | --- |
|  | Patients (%)<br>n = 63 | Number of<br>ADRs (%)<br>n = 84 | Patients (%)<br>n = 64 | Number of ADRs<br>(%)<br>n = 131 |
| **Body as a whole** | **1 (1.6)** | **1 (1.2)** | **4 (6.2)** | **4 (3.1)** |
| Fever | 1 (1.6) | 1 (1.2) | 2 (3.1) | 2 (1.5) |
| Asthenia | - | - | 2 (3.1) | 2 (1.5) |
| **Cardiovascular** | **-** | **-** | **2 (3.1)** | **2 (1.5)** |
| Syncope | - | - | 1 (1.6) | 1 (0.8) |
| Tachycardia | - | - | 1 (1.6) | 1 (0.8) |
| **Digestive** | **11 (17.5)** | **12 (14.3)** | **21 (32.8)** | **26 (19.8)** |
| Vomiting | 8 (12.7) | 8 (9.5) | 4 (6.2) | 4 (3.1) |
| Nausea | 2 (3.2) | 2 (2.4) | 8 (12.5) | 8 (6.1) |
| Dyspepsia | 1 (1.6) | 1 (1.2) | 8 (12.5) | 8 (6.1) |
| Abdominal pain | 1 (1.6) | 1 (1.2) | 4 (6.2) | 4 (3.1) |
| Others | - | - | 2 (3.1) | 2 (1.5) |
| **Hematological** | **6 (9.5)** | **7 (8.3)** | **-** | **-** |
| Eosinophilia | 3 (4.8) | 3 (3.6) | - | - |
| Leukopenia | 3 (4.8) | 3 (3.6) | - | - |
| Plaquetopenia | 1 (1.6) | 1 (1.2) | - | - |
| **Nutritional** | **30 (47.6)** | **31 (36.9)** | **40 (62.5)** | **44 (33.6)** |
| Weight loss | 13 (20.6) | 13 (15.5) | 33 (51.6) | 33 (25.2) |
| Hyporexia | 18 (28.6) | 18 (21.4) | 11 (17.2) | 11 (8.4) |
| **Musculoskeletal** | **1 (1.6)** | **1 (1.2)** | **2 (3.1)** | **2 (1.5)** |
| Myalgias | - | - | 2 (3.1) | 2 (1.5) |
| Chest pain | 1 (1.6) | 1 (1.2) | - | - |
| **CNS** | **19 (30.2)** | **22 (26.2)** | **28 (43.8)** | **39 (29.8)** |
| Headache | 6 (9.5) | 6 (7.1) | 20 (31.2) | 20 (15.3) |
| Irritability | 15 (23.8) | 15 (17.9) | 7 (10.9) | 7 (5.3) |
| Dizziness | - | - | 5 (7.8) | 5 (3.8) |
| Others | 1 (1.6) | 1 (1.2) | 5 (7.8) | 7 (5.3) |
| **Respiratory** | **1 (1.6)** | **3 (3.6)** | - | - |
| Acute bronchitis | 1 (1.6) | 1 (1.2) | - | - |
| Rhinorrhea | 1 (1.6) | 1 (1.2) | - | - |
| Influenza Syndrome | 1 (1.6) | 1 (1.2) | - | - |
| **Skin** | **6 (9.5)** | **7 (8.3)** | **10 (15.6)** | **13 (9.9)** |
| Rash | 6 (9.5) | 7 (8.3) | 6 (9.4) | 6 (4.6) |
| Urticaria | - | - | 4 (6.2) | 4 (3.1) |
| Others | - | - | 3 (4.7) | 3 (2.3) |
| **Psychiatric** | - | - | **1 (1.6)** | **1 (0.8)** |
| Depression | - | - | 1 (1.6) | 1 (0.8) |
| **Total** | **63 (100.0)** | **84 (100.0)** | **64 (100.0)** | **131 (100.0)** |

Detailed description of the 215 ADRs occurring in the 127 patients segregated by organ system. For each age group, two columns are shown. The first column, *Patients with ADRs*, shows the number of patients presenting at least one ADR and its corresponding percentage. The second column, *Number of ADRs*, depicted the observed number of ADRs and its corresponding percentage. Low frequency symptoms were grouped into a general category, namely, "*others*". *digestive: (epigastralgia; pyrrhosis)*. *CNS:(depression; nightmare; dysarthria; insomnia; loss of memory; panic attacks; tinnitus)*. *Skin: (facial edema; pruritus)*.

ADRs had an earlier onset in adults, who presented a median onset time of 6.5 days (IQR_25-75_ = 1.5 - 9), compared to children, who presented a median onset time of 12 days (IQR_25-75_ = 9.7 - 21; W.T P = 2.9. 10^-4^).

### Severity

ADRs severity is described in Table 2. Most ADRs were mild (74.9%) and moderate (16.3%) and resolved without sequelae. Severe ADRs were infrequent: 2/215 (0.9%). Severe ADRs occurred in 1 adult and in 1 child (see Table 2). The adult was a woman in her 30’s, who developed a headache with a defined relationship to NF. ADR resolved without consequences, but NF treatment was discontinued. The child was in the 8mos-2yrs age range, who presented severe leukopenia, but also resolved without consequences and he was able to complete treatment.

The severity of ADRs was associated with treatment discontinuation in adults, with 10/15 (67%) of discontinuations in subjects that presented moderate/severe ADRs and 3/49 (6.1%) in subjects that presented mild ADRs (F.E.T, OR = 27.8, CI95% = [5.1 - 212]; P_Adjusted_ = 1.6.10^-5^). In children no association was observed (F.E.T, OR = 2.4, CI95% = [0.04 - 35]; P_Adjusted_ = 0.4).

Serious events were observed in 2 patients. One patient who died was in the 0-8 month age range, coinfected with *T.cruzi* and HIV by the transplacental route as mentioned above (NF unrelated serious adverse event) and the other patient a female in her 30’s who presented tremors, dysarthria and panic attacks which required hospitalization. All ADRs were resolved without consequences but treatment was discontinued.

### Treatment completion

Overall treatment was completed in 289/320 patients (90.3%) without differences between children (92.6%) and adults (85.7%; Table 4). Treatment discontinuation took place in 31/320 (9.7%) patients, but only 14/320 (4.4%) were related to ADRs (Table 5). ADRs-related discontinuations occurred more frequently in adults (9.5%) than in children (1.9%; F.E.T, OR = 5.5, CI95% = [1.5 - 24]; P_Adjusted_ = 0.008). Notably, the main cause of treatment discontinuation in children was related to moderate skin ADRs (3/4 children).

**Table 4.** Treatment discontinuation and interruption.

|  | Children (%) | Adults (%) | Total Patients (%) |
| --- | --- | --- | --- |
| **Complete treatment** |  |  |  |
| Yes | 199 (92.6) | 90 (85.7) | 289 (90.3) |
| No | 16 (7.4) | 15 (14.3) | 31 (9.7) |
| **Discontinuation cause** |  |  |  |
| Patient Decision | 6 (2.8) | 1 (1.0) | 7 (2.2) |
| Adverse Effect | 4 (1.9) | 10 (9.5) | 14 (4.4) |
| Death | 1 (0.5) | - | 1 (0.3) |
| Lost of follow-up | 5 (2.3) | 4 (3.8) | 9 (2.8) |
| Treatment Complete | 199 (92.6) | 90 (85.7) | 289 (90.3) |
| **Temporary Interruption** |  |  |  |
| Yes | 10 (4.7) | 10 (9.5%) | 20 (6.2) |
| No | 205 (95.3) | 95 (90.5) | 300 (93.8) |
| **Total** | **215 (67.0)** | **105 (33.0)** | **320(100.0)** |

Detailed description of reasons of treatment discontinuation and interruption for all patients included in this study (n = 320 patients).

**Table 5.** ADRs causing treatment discontinuation.

|  | Age range (yrs.) | Gender | Symptoms | Treatment Length (days) | Second Treatment |
| --- | --- | --- | --- | --- | --- |
| **Pediatrics** | 0-1 | F | Irritability | 15 | NF, Completed |
|  | 7-17 | M | Rash | 15 | - |
|  | 7-17 | F | Rash | 27 | - |
|  | 2-6 | M | Abdominal pain, Rash, Fever, Eosinophilia | 21 | - |
| **Adults** | 30-39 | F | Headache, Irritability, Nausea | 3 | BZ, discontinued because of ADR |
|  | 40-49 | F | Hiporexia | 23 | BZ, Completed |
|  | 50-59 | F | Hiporexia | 11 | BZ, Completed |
|  | 30-39 | F | Syncope, Dizziness, Headache, Dyspepsia, Nausea | 12 | BZ, Completed |
|  | 40-49 | F | Headache, Abdominal pain | 15 | - |
|  | 20-29 | F | Abdominal pain, Vomiting, Myalgias | 11 | - |
|  | 30-39 | F | weight loss, Irritability, tremors, dysarthria, Panic attack | 18 | BZ, discontinued because of ADR |
|  | 20-29 | F | Rash, Itching, Headache, Dyspepsia | 12 | BZ, Completed |
|  | 30-39 | F | Headache. Dizziness | 8 | BZ, Completed |
|  | 30-39 | F | Psychomotor agitation | 10 | BZ, discontinued because of ADR |

Detailed ADR description for those patients who discontinued treatment due to ADRs. NF: nifurtimox, BZ: benznidazole. All patients had a good response to symptomatic treatment.

A total of 20/320 (6.2%) subjects temporally interrupted NF (7 adults and 5 children due to ADRs and 8 by patient decision), without differences among adults (9.5%) and children (4.7%, F.E.T, OR = 2.1, CI95% = [0.77 - 5.9]; P_Adjusted_ = 0.14). The median temporary interruption length was 7 days (IQR_25-75_: 2-9 days) with no differences between adults and children.

ADRs leading to temporary treatment interruptions were digestive (5), CNS (3), skin (2), cardiovascular (2), and nutritional (2) among adults, and digestive (2), skin (2) and hematologic (2) among children. Treatment discontinuation occurred in 4 of these 12 patients (2 adults, 2 children). The remaining patients completely recovered after symptomatic treatment and/or transient interruption of NF.

For those 289 patients who completed treatment, mean dose, number of tablets and the length of treatment are described in Table 6.

**Table 6:**
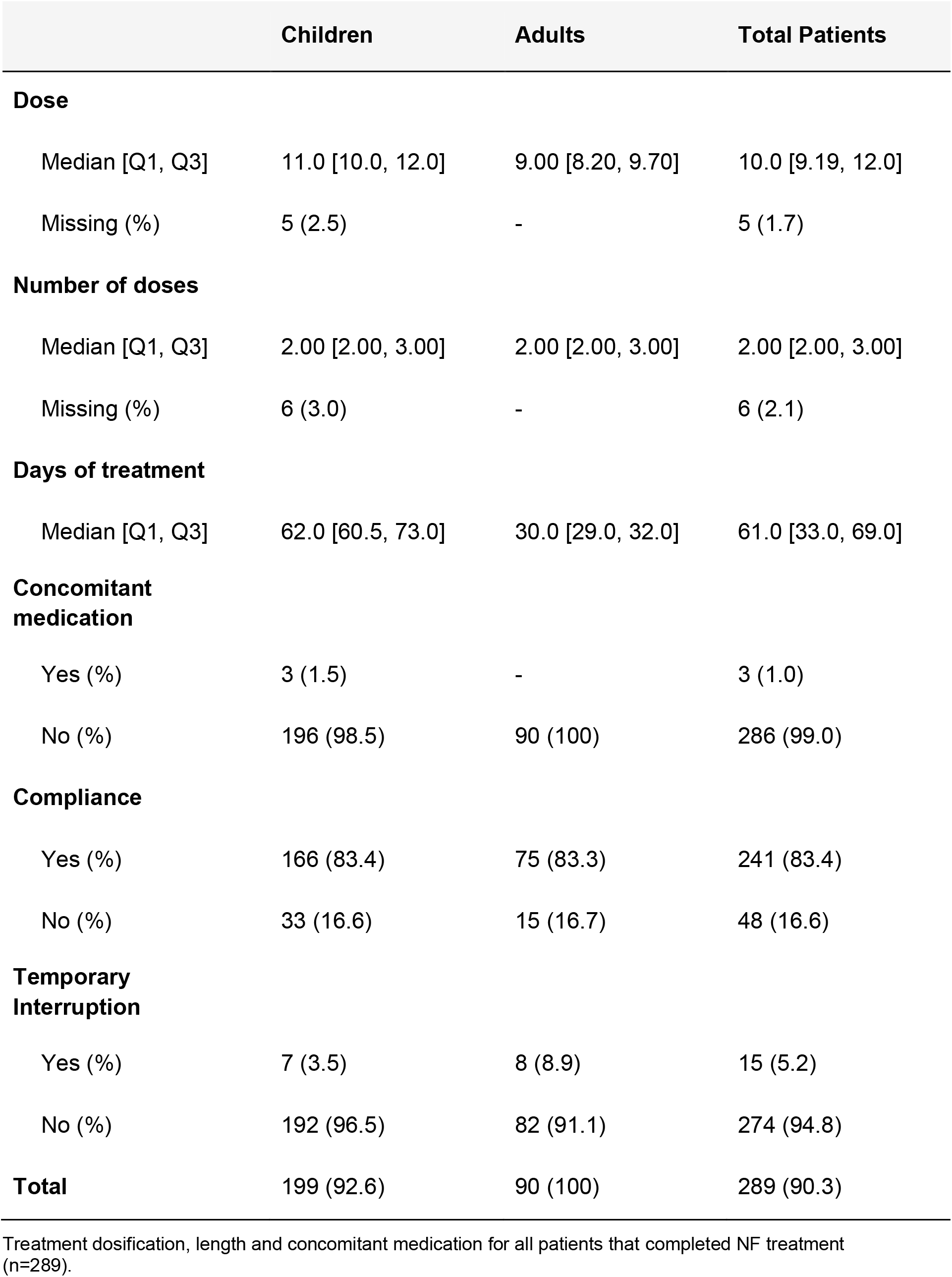
Treatment description.

|  | Children | Adults | Total Patients |
| --- | --- | --- | --- |
| **Dose** |  |  |  |
| Median [Q1, Q3] | 11.0 [10.0, 12.0] | 9.00 [8.20, 9.70] | 10.0 [9.19, 12.0] |
| Missing (%) | 5 (2.5) | - | 5 (1.7) |
| **Number of doses** |  |  |  |
| Median [Q1, Q3] | 2.00 [2.00, 3.00] | 2.00 [2.00, 3.00] | 2.00 [2.00, 3.00] |
| Missing (%) | 6 (3.0) | - | 6 (2.1) |
| **Days of treatment** |  |  |  |
| Median [Q1, Q3] | 62.0 [60.5, 73.0] | 30.0 [29.0, 32.0] | 61.0 [33.0, 69.0] |
| **Concomitant medication** |  |  |  |
| Yes (%) | 3 (1.5) | - | 3 (1.0) |
| No (%) | 196 (98.5) | 90 (100) | 286 (99.0) |
| **Compliance** |  |  |  |
| Yes (%) | 166 (83.4) | 75 (83.3) | 241 (83.4) |
| No (%) | 33 (16.6) | 15 (16.7) | 48 (16.6) |
| **Temporary Interruption** |  |  |  |
| Yes (%) | 7 (3.5) | 8 (8.9) | 15 (5.2) |
| No (%) | 192 (96.5) | 82 (91.1) | 274 (94.8) |
| **Total** | 199 (92.6) | 90 (100) | 289 (90.3) |

Treatment dosification, length and concomitant medication for all patients that completed NF treatment (n=289).

### Pediatric cohort analysis

A sub-analysis by age group of the pediatric cohort was carried out to elucidate whether there was any trend in the number, frequency or type of ADRs.

A high rate of treatment completion (>89%) was observed without differences within pediatric age groups.

No significant differences among pediatric subgroups were observed in the rates of temporary interruption (lower than 5% in all pediatric groups) or in the rate of ADRs (25-38%). In addition, a high compliance (greater than 70%) was observed in all groups and notably > 90%) in patients under 2 years old (Supplementary Table 1).

The most frequently observed pediatric ADRs were nutritional, followed by CNS adverse reactions, without clear differences among age groups. Headache was most frequent in children older than 7 years of age. Digestive and hematological events were mainly observed in children younger than 2 years old (Table 7).

**Table 7:**
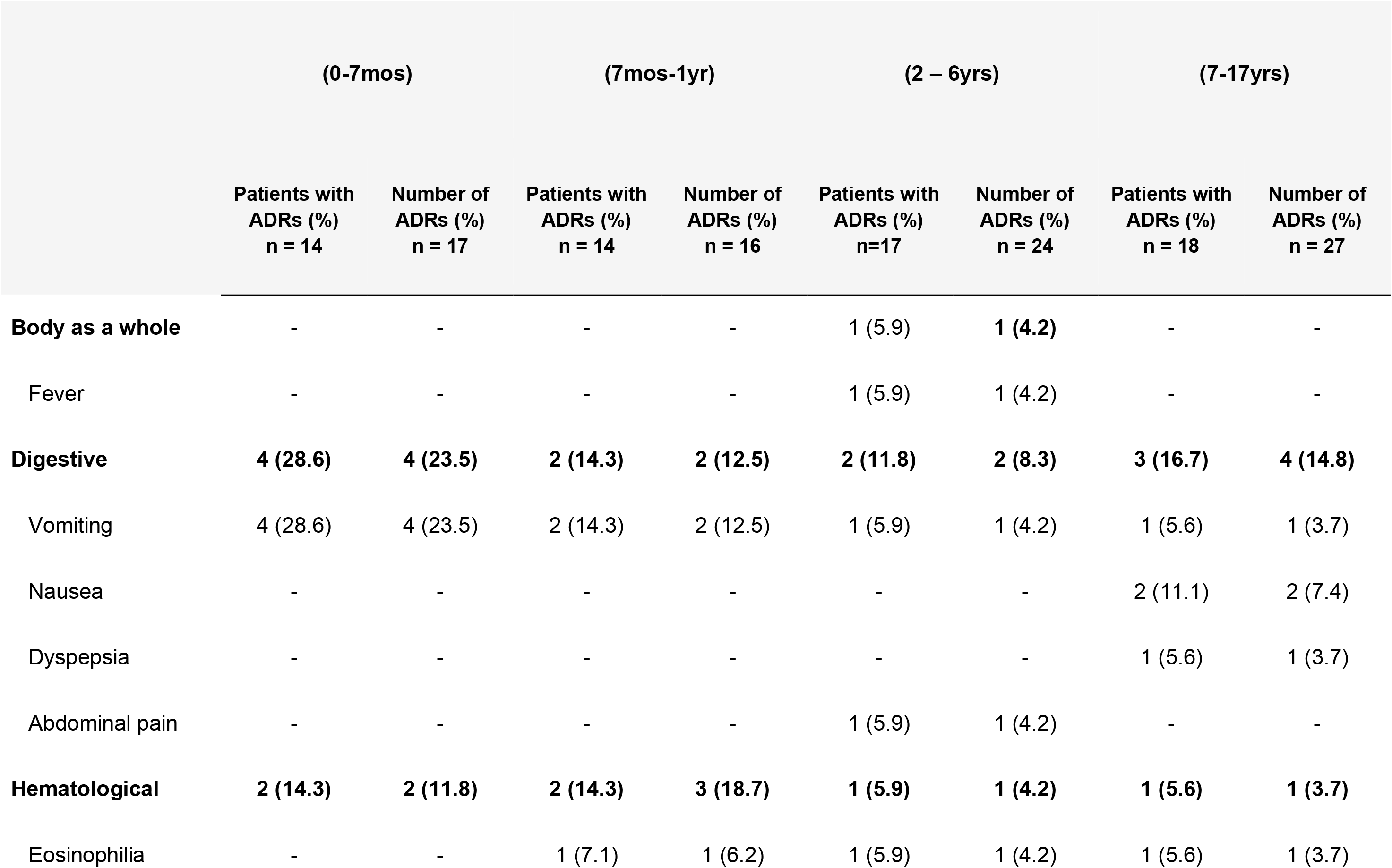
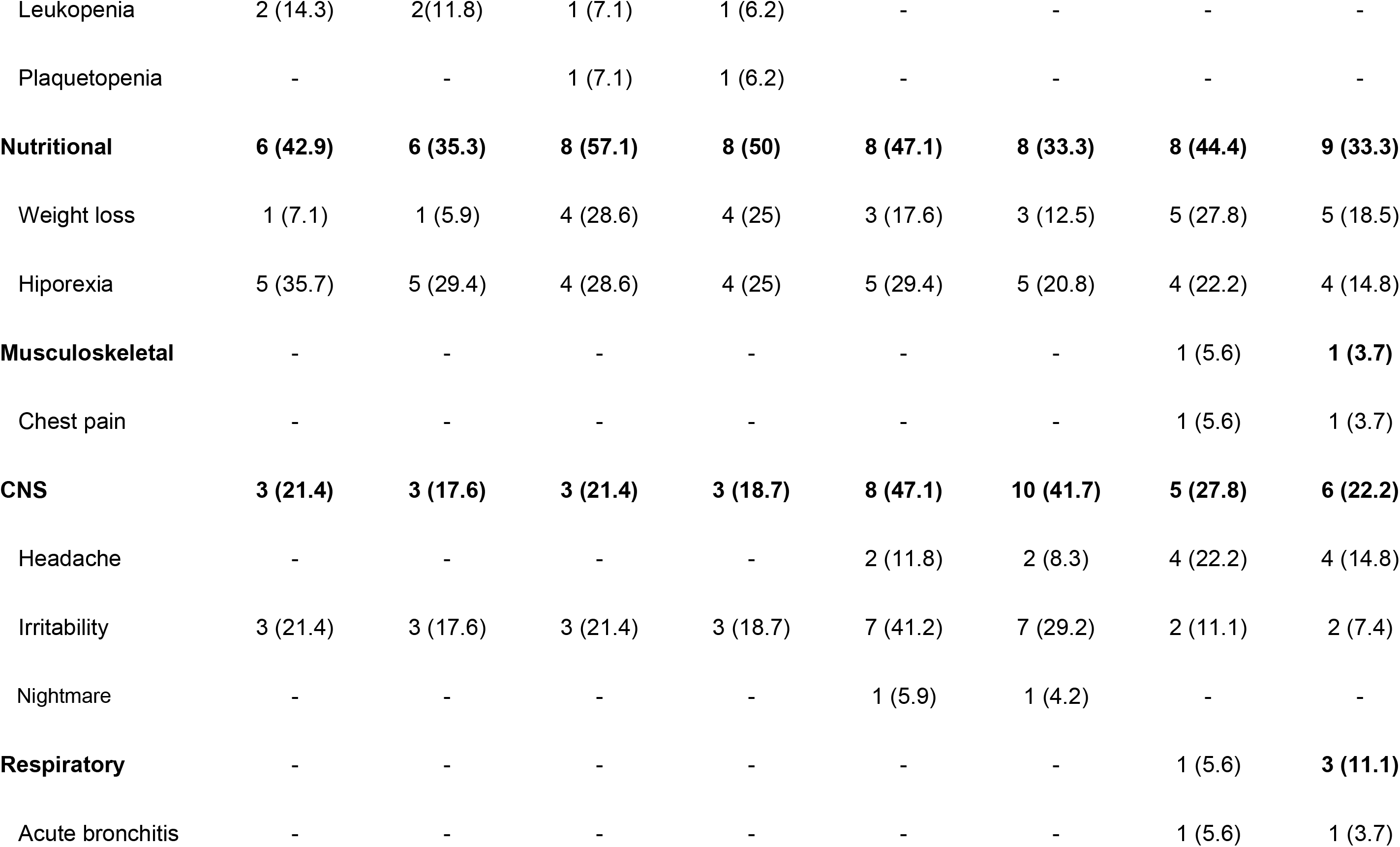
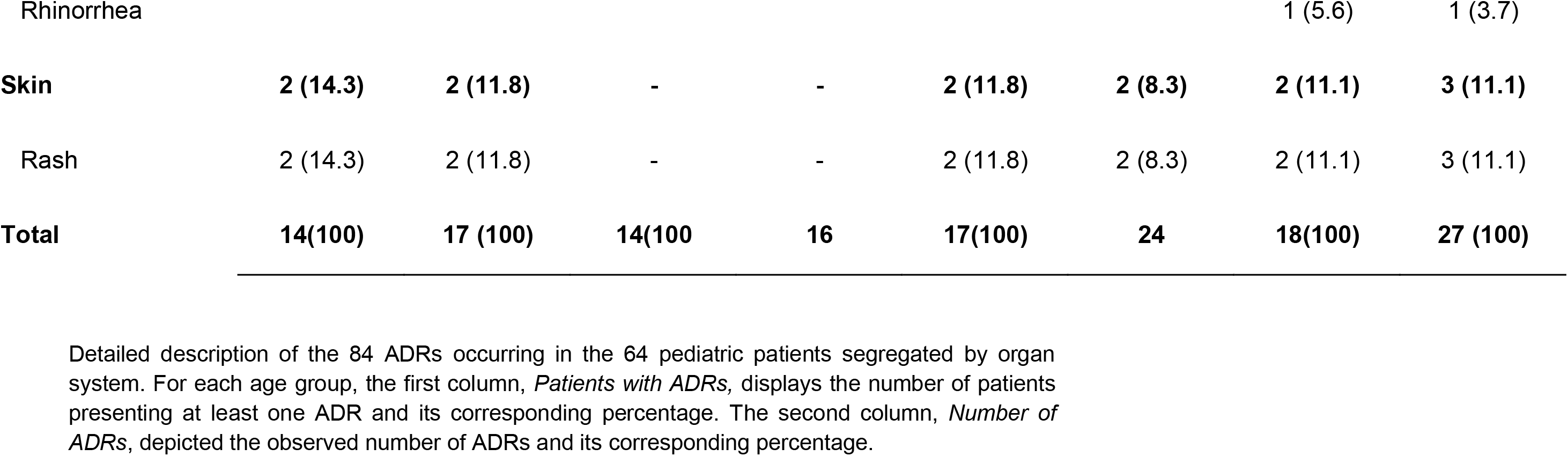
ADR occurrence and patient incidence by organ system.

## Discussion

We present a large cohort of ChD patients (children and adults), treated with NF. We observed a low loss to follow-up, similar to other prospective pediatric and adult ChD studies [11–14]. ADRs incidence in our study was strongly associated with patient age, since adults had higher incidence of ADRs and related treatment discontinuations than children. Moreover, NF-related ADRs were significantly more frequent in adults. Although pediatric pharmacologic studies on NF are still lacking, it is possible that observed differences in incidence of ADRs among the pediatric and adult sub-populations could be due to age-related differences in drug metabolism. NF is metabolized in the liver, and, similar to many other drugs [15–17], it would be expected to undergo faster liver clearance in children compared to adults, leading in shorter half-lives and steady-state plasma concentrations. Pharmacological NF studies are currently underway to clarify this issue (www.clinicaltrials.gov NCT01927224, NCT02625974).

NF related ADRs had a lower incidence in our study compared to previous reports [4,11–13]. Moreover, the rate of ADRs per patient in our adult cohort was 4-fold lower than the reported in previous studies [13]. These differences could be explained by a larger sample size of our cohort that would yield a more accurate estimate of the incidence of ADRs. However, some alternative explanations are possible, such as socio-economic or ethnical differences among the studied populations, detection bias (our data were retrospectively collected for this analysis), or other yet unknown issues.

The ADRs observed in our cohort were mostly mild. Severe events were observed in 3.2% of patients and all patients recovered with no sequelae. This is comparable to previous studies in adults [13]. However, a greater incidence (7,4%) of serious adverse events was reported by Jackson et.al.[4].

ADRs appeared earlier in adults than in children but most of them occurred within the first month of treatment, suggesting that most NF ADRs are not dependent on cumulative doses.

ADRs profiles were similar between pediatric and adult populations, except for hematological ADRs, which appeared only in children, mainly those under 2 years of age. The most frequent ADRs were nutritional, mainly hyporexia and weight loss, in line with the observations of many other ChD researchers [4,11,13,14]

CNS ADRs are a major concern. Seizures and psychiatric ADRs related to NF have been previously described [18]. NF has a high level of fat solubility and is well distributed throughout the tissues, including the CNS. Even though NF is a substrate of breast cancer resistance protein (BCRP), which may be responsible for the active transfer of NF out of the CNS, it is possible that some patients may have BCRP polymorphisms that decrease this transfer, thus exposing them to high level of NF in the CNS for longer periods of time (i.e. exposing them to CNS ADRs) [19,20]. However, this remains a hypothesis to be tested. We observed mostly headaches and CNS irritability as NF ADRs [19]. An adult patient presented tremors, dysarthria and panic attacks which required hospitalization, but recovered without consequences after temporary treatment interruption. Evaluation of CNS ADRs such as headaches, was difficult in children and particularly in infants because only older children can accurately express these symptoms. However, associated signs, such as unexplained irritability, food refusal, or vomiting, were not reported by caregivers or observed by our pediatricians who have significant experience in evaluating ChD pediatric patients.

The profile of digestive ADRs was similar to that reported in other NF studies [4,5,12,13,21]. In children, digestive ADRs could be related to the lack of an appropriate pediatric formulation of NF which requires pill fractioning. As pill fragments are not easily (or willingly) swallowed by small children, this sometimes results in vomiting and other problems that may not be specifically related to the active drug. A new pediatric NF formulation in the late stages of clinical development (clinicaltrials.gov NCT02625974), over time, would eventually be helpful to address the pediatric formulation gap and possibly decrease the incidence of digestive ADRs.

Skin reactions are the main ADRs observed during treatment with the alternative drug BZ [6], but are much less frequently described with NF. Accordingly, we observed few skin manifestations in our cohort, and only 3 children developed skin reactions that led to treatment discontinuation. The differences in ADR profiles between BZ and NF are not clearly explained to date, particularly given that they are both nitro-drugs. Unfortunately, NF metabolism, and metabolite profiles for both drugs remain poorly studied, which hampers any speculation on the pharmacological reasons behind these ADR differences [22,23].

Pediatric treatment discontinuation rates due to NF ADRs in our study were comparable with those in other studies[11,12]. Even though we observed higher discontinuation rates in adults compared to children, these adult rates (i.e. 14.3%) were lower than those reported in previous studies, which ranged from 19.8 % to 43.8% [4,13]. This difference could be due to the fact that most of the adult patients in our study were relatives of previously treated and cured children, to whom we offered treatment as part of our ChD family screening and treatment protocol. This population is highly motivated to persist and complete treatment.

In summary, our results suggest that NF is a safe drug to use in both pediatric and adult ChD patients. Since more primary infections of ChD occur during childhood, early diagnosis and treatment of children is vital to prevent long-term ChD sequelae. Under the light of the findings supporting NF safety in childhood, we believe that treatment should not be delayed.

Our study supports that NF is a safe drug for ChD treatment at any age since most treated patients only showed mild ADRs. Infants and toddlers seem to be at very low risk for severe ADRs and there is no reason to postpone treatment based on the theoretical risk.

## Data Availability

Not applicable

## Financial support

This work was supported by an Institution initiative research by Bayer [grant number PR5071885]. A.J.B., F.G.B., G.M. and J.A, are researchers in the National Scientific and Technical Research Council of Argentina (CONICET).

## Potential conflicts of interest

JA is a consultant of Bayer. All other authors report no potential conflicts. All authors have submitted the ICMJE Form for Disclosure of Potential Conflicts of Interest

## Supplementary Material

Supplementary Table 1: Treatment completion, compliance, interruption and discontinuation causes for different age groups in the pediatric cohort. Also, the number and rates of adverse events on different age groups in the pediatric cohort are shown.

Article’s main point

This work shows that children were less affected than adults by ADRs associated with nifurtimox treatment. All the evidence, the incidence, the rate of ADRs and discontinuations suggest that treatment should not be delayed for safety concerns.

## Notes

### Clinical Trial

NCT04274101

### Author Declarations

Study protocol was approved by the research and teaching committee and the bioethics committee of the Children Hospital Ricardo Gutierrez, Buenos Aires.

